# A Simple Method to Estimate Flow Restriction for Dual Ventilation of Dissimilar Patients: The BathRC Model

**DOI:** 10.1101/2020.04.12.20062497

**Authors:** Andrew Plummer, Jonathan L. du Bois, Joseph Flynn, Jens Roesner, Siu Man Lee, Patrick Magee, Malcolm Thornton, Andrew Padkin, Harinderjit S. Gill

**Affiliations:** Department of Mechanical Engineering, University of Bath, UK; Royal United Hospitals NHS Foundation Trust, Bath, UK; BMI Bath Clinic, Bath, UK; Centre for Therapeutic Innovation, University of Bath, UK

**Author notes:** corresponding author, address: Dept. of Mechanical Engineering, University of Bath, Claverton Down, Bath, BA2 7AY. Author email addresses.

**Keywords:** Acute Respiratory Distress Syndrome, Coronavirus, COVID-19, Differential multi-ventilation, Dual ventilation, Patient matching, Ventilator sharing, Ventilator splitting

## Abstract

**Objectives:** With large numbers of COVID-19 patients requiring mechanical ventilation and ventilators possibly being in short supply, in extremis two patients may have to share one ventilator. Careful matching of patient ventilation requirements is necessary. However, good matching is difficult to achieve as lung characteristics can have a wide range and may vary over time. Adding flow restriction to the flow path between ventilator and patient gives the opportunity to control the airway pressure and hence flow and volume individually for each patient. This study aimed to create and validate a simple model for calculating required flow restriction.

**Design/Setting:** Experimental/Secondary Care

**Methods:** We created a simple linear resistance-compliance model, termed the BathRC model, of the ventilator tubing system and lung allowing direct calculation of the relationships between pressures, volumes, and required flow restriction. Experimental measurements were made for parameter determination and validation using a clinical ventilator connected to two test lungs. For validation, differing amounts of restriction were introduced into the ventilator circuit.

**Results:** The BathRC model was able to predict tidal lung volumes with a mean error of 4% (min:1.2%, max:9.3%).

**Conclusion:** We present a simple model validated model that can be used to estimate required flow restriction for dual patient ventilation. The BathRC model is freely available; this tool is provided to demonstrate that flow restriction can be readily estimated.

**ARTICLE SUMMARY:** Strengths and limitations of this study

- Data collected on a clinical system
- Simple model, easy to implement
- Validation performed
- Limited to 2 types of restrictors tested
- Data from only one ventilator type

## INTRODUCTION

The current COVID-19 crisis could risk ventilator capacity shortfall. If necessary, ventilator capacity could be increased by ventilating two patients using the machine, namely Dual Patient Ventilation (DPV). This would require two circle systems, connected in parallel, as per the work of Neyman & Irvin^1^, and Paladina et al.^2^. Complexity arises when patients have different airway and lung impedances as each would require different ventilator settings. Hence, distributing and controlling pressure and flow to each patient, independently, is a significant challenge. Accordingly, the Anesthetic Patient Safety Foundation (APSF) has mandated against ventilator sharing^3^.

The COVID-19 crisis has renewed interest in DPV and, in March 2020, this approach was introduced in New York, adopting the “Columbia Protocol” of ventilation^4^. This relies on careful matching of patient characteristics, and its effectiveness would be sensitive to changes in patient compliance. A new experimental study by Tronstad et al.^5^in relation to COVID-19 concluded that large discrepancies were found in delivered tidal volumes for paired test lungs with compliance differences. Furthermore, high Positive End-Expiratory Pressure (PEEP) could strongly influence the distribution of tidal volume. They were unable to reliably overcome this hazard.

Evidence is emerging that lung compliance in COVID-19 patients is not as reduced as in other forms of Acute Respiratory Distress Syndrome, ARDS^6^. Nevertheless, DPV for patients with differing characteristics, particularly tidal volume due to differing compliances, will still require a modified breathing circle. This refers to the introduction of an impedance (resistance or compliance) to appropriately distribute the supplied tidal volume. Increasing the resistance in the inspiratory limb of the patient with either the higher lung compliance or the one requiring a lower tidal volume seems plausible. This is the subject of this paper. A team from Hospital Geel, Belgium, have been experimenting with the same technique^7^, and there has been a recent simulation study^8^. This latter work succinctly reviews previous work on DPV and the challenges involved.

A single circle system for one patient has a unidirectional valve in each of its inspiratory and expiratory limbs. These are usually integrated into the anaesthetic machine circle system attachments. A key risk of DPV is inadvertent sharing of gas flows either between patients or between the inspiratory and expiratory limbs of a single patient. Installing two parallel circle systems, each with two directional control valves, restores some control over this risk. This arrangement reduces dead space for each half of the system, and the potential for CO_2_ rebreathing. The testing reported here does not include studies of how the arrangement manages CO_2_.

Mathematical modelling and simulation of both human respiratory and mechanical ventilation systems is invaluable to help understand novel scenarios such as DPV. Characterising lung mechanical properties using resistance and compliance has become commonplace. Estimated values are available from studies such as Arnal et al.^9^, although other modelling approaches are possible as reviewed in Carvalho and Zin,^10^. Complete system models have also been developed as reported in Wilson et al.,^11^ and these have been used to extensively study low flow breathing systems^12^. However, the aim of our study was to determine if a very simple analytical model can adequately predict behaviour of a ventilator system. An analytical model permits direct calculation of the flow restrictor resistance required to achieve a specified tidal volume.

## METHODS

### A linear lumped resistance-compliance (RC) network model: the BathRC model

A highly simplified lumped resistance-compliance model (Figure 1) can be used to represent single patient ventilation with four terms: linear resistance (*R_v_*) and compliance (*C_v_*) for the ventilator tubing system, and linear resistance (*R*) and compliance (*C*) for the patient. The inspiration and expiration phases can be represented with different parameters, resulting in up to eight parameters per patient. This RC model calculates tidal volume as a function of variations in patient *R* and *C* values. It also gives the relationships between airway flow rate, q, airway pressure, *p_p_*, and ventilator pressure, *p* (full equations in supplementary information, Appendix A). A DPV-specific use case is to calculate the required inspiration restriction (*R_r_*) to operate with an increased ventilator pressure (necessitated by ventilator sharing) without increasing the patient’s tidal volume undesirably. Experimental measurements are required for model parameter estimation and to validate the model. The experiments are described below.

**Figure 1.**
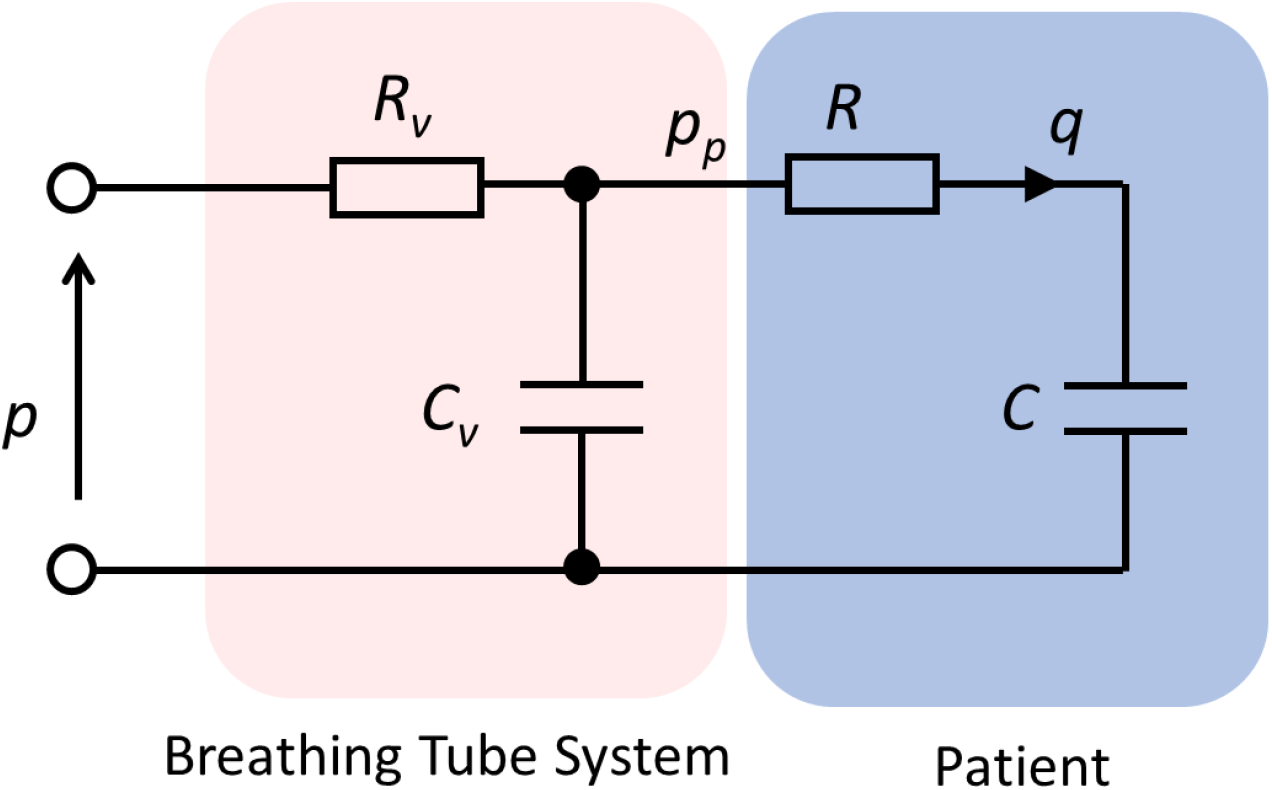
Simple RC network model of ventilator system and patient, with linear resistance (*R_v_*) and compliance (*C_v_*) for the ventilator tubing system, and linear resistance (R) and compliance (C) for the patient. The RC model calculates tidal volume as a function of variations in patient *R* and *C* values. It also gives the relationships between airway flow rate, q, airway pressure, *p_p_*, and ventilator pressure, *p*.

### Experimental Measurements

Experiments were performed using an Aisys CS^2^ (Software version 8.0, GE Healthcare, Chicago, USA) anaesthetic ventilator operated in Pressure Control Mode. This is the most appropriate setting for DPV, as the settings for a single patient would not need to change for ventilating two identical patients. In this mode the adjustable settings are:

- Pinsp (the inspiration pressure in excess of PEEP).
- PEEP (Positive end-expiratory pressure), the ventilator pressure during expiration.
- RR (the respiratory rate, breaths per minute).
- I:E (inspiration to expiration time ratio).

In Pressure Control Mode, the ventilator effectively controls the driving pressure to transition between PEEP and Pinsp+PEEP as quickly as possible at the required switching times; any limits or triggers which might alter this profile need to be disabled for dual patient use.

The ventilator was connected to a Silverknight 22 mm circle system (Intersurgical Ltd, Wokingham, UK) in conjunction with Heat and Moisture Exchange (HME) filters (Clear-Therm 3, Intersurgical Ltd). Two fixed test lungs (Test Lung 190, Siemens Healthineers, Erlangen, Germany) were used for the experiments, these were termed *Lung 1* and *Lung 2*. Two Fluke VT Plus HF Gas Flow Analysers (Fluke Biomedical, Everett, Washington, USA) were used to make flow measurements, and data were collected by connecting each analyser to a personal computer (Dell XPS13 i5, Dell UK, Bracknell, UK) running Vent Tester for Windows software (version 2.01.07, Fluke Biomedical). Data were collected at 50 Hz on each personal computer. Custom functions (MATLAB 2019b, The Mathworks Inc., Natick, MA, USA) were used to co-register the data collected on the two computers for each experiment.

Figure 2 details three configurations that were used for experimental validation of the model. The characterisation experiments used single circuit configurations (Figures 2a, Circuit 1, and 2b, Circuit 2) and validation measurements used dual circuit configurations (Figure 2c, Circuit 3). The dual circuit layout contained four sets of non-return valves (Ref: 1950000, Intersurgical Ltd), also known as one-way valves to prevent sharing of gas flows and to handle expired CO_2_ adequately; the proposed circuit (Figure 2c) used additional non-return valves in each inspiratory and expiratory limb to stop inspiration or expiration back flows and to reduce each system’s dead space.

**Figure 2.**
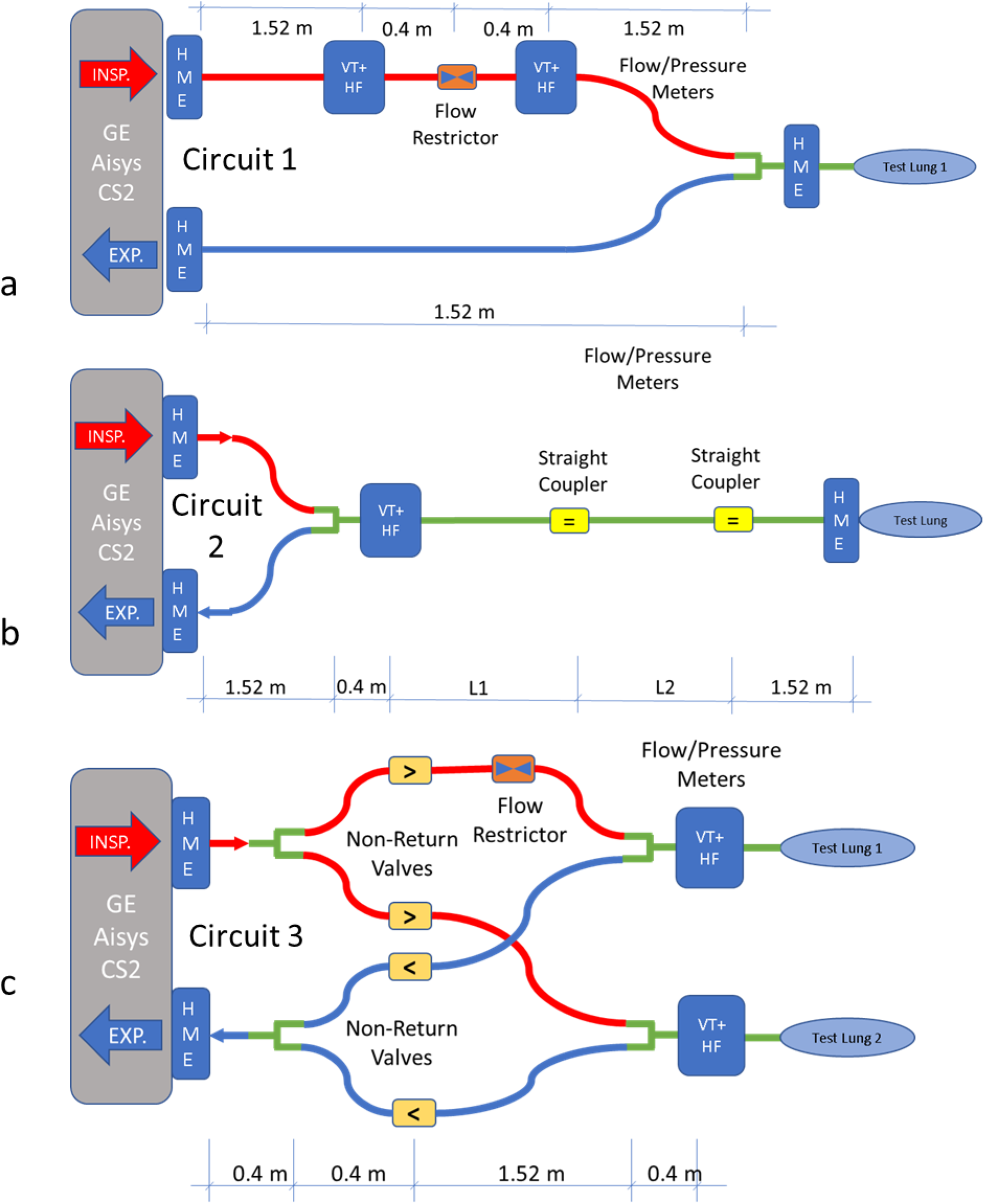
Ventilators circuits used for testing and validation, HME=heat and moisture exchanger, VT+ HF= Fluke VT+ HF flow meter, INSP.=inspiration, EXP. = expiration.

### Parameter Estimation

Circuit 1 was used to estimate flow resistance parameters for different components. Two types of restrictor were tested, the first was a non-return valve and the second was a novel flow restrictor with a very small orifice (11.7 mm^2^ effective cross-section), hereafter termed *small orifice restrictor* or *SOR*. The SOR device was 3D printed (Form 2, Formlabs Inc., Somerville, MA, USA) for testing purposes. A minimum of 15 cycles of data were collected at each of three different Pinsp pressures, 5, 15, 25 cmH_2_O; for all tests PEEP was set to 5 cmH_2_O, RR was 15 breaths/min, and I:E ratio was 1:2. The use of the two flow meters in Circuit 1 were used to measure the pressure drop (∆*p*) across each tested component. For each flow restrictor, all pressure drop measurements were plotted against the mean flowrate (*Q_m_*), the average of the flowrates measured from the two flowmeters. A bi-square weighted robust least squares fitting method (Matlab 2019b, The MathWorks, Natick, MA, USA) was used to fit the quadratic function,

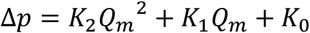

This equation becomes linear by setting *K_2_=0*, and proportional by additionally setting *K_0_=0*. Hence, the differences between quadratic, linear and constrained (proportional) linear fits were examined. With a proportional fit, *K_1_* is the resistance value of the component being tested. The BathRC model formulation can only accommodate the proportional linear representation of a restrictor.

Circuit 2 was used to estimate the compliance and resistance values for both test lungs (each lung was tested separately). Here, L1=L2=0.4 m to give a total tube length of 2.32 m beyond the flow meter. The compliance of the tubing itself was estimated using L1=L2=1.52 m, to make the total length 4.56 m, and replacing the test lung with a blockage at the end of the tube.

For all three tests, at least 15 cycles were again collected at each of three different Pinsp pressures, 5, 15, 25 cmH_2_O, keeping the values of PEEP, RR and I:E fixed as above.

### Validation of BathRC Model

Circuit 3 was used for validation and data were collected for the Pinsp pressures of 15 and 25 cmH_2_O (other ventilator settings were fixed). The following five validation tests were conducted with different flow restrictors connected in the inspiratory limb for Lung 1 (Figure 2c):

1. Pinsp = 25 cmH_2_O and no restrictor present (replaced by straight connector)
2. Pinsp = 25 cmH_2_O and non-return valve acting as restrictor
3. Pinsp = 25 cmH_2_O and SOR device
4. Pinsp = 15 cmH_2_O and no restrictor present (replaced by straight connector)
5. Pinsp = 15 cmH_2_O and non-return valve acting as restrictor

The measured values of tidal volume (obtained from the integration of the measured flows) were compared predictions given by the BathRC model and estimated parameters (as above).

## RESULTS

### Parameter Estimation

Figure 3 a shows the two linear fits for pressure drop versus mean flowrate (Circuit 1 tests) in the case of the non-return valve. The corresponding fit data is presented in Table 1, with R^2^ values of 0.99 and 0.75 for linear and proportional fits, respectively. The data show that this non-return valve has a cracking pressure of approximately 1 cmH_2_O. For the SOR, the quadratic and constrained linear fits are shown in Figure 3 b. The fits (Table 1) result in R^2^ values of 0.99 and 0.89 for quadratic and proportional fits, respectively. Resistances for the two devices were estimated from the gradient of the proportional fits. The non-return valve resistance, *R_r_*, was 12 cmH2O/L/s, and the SOR resistance was 33 cmH2O/L/s.

**Figure 3.**
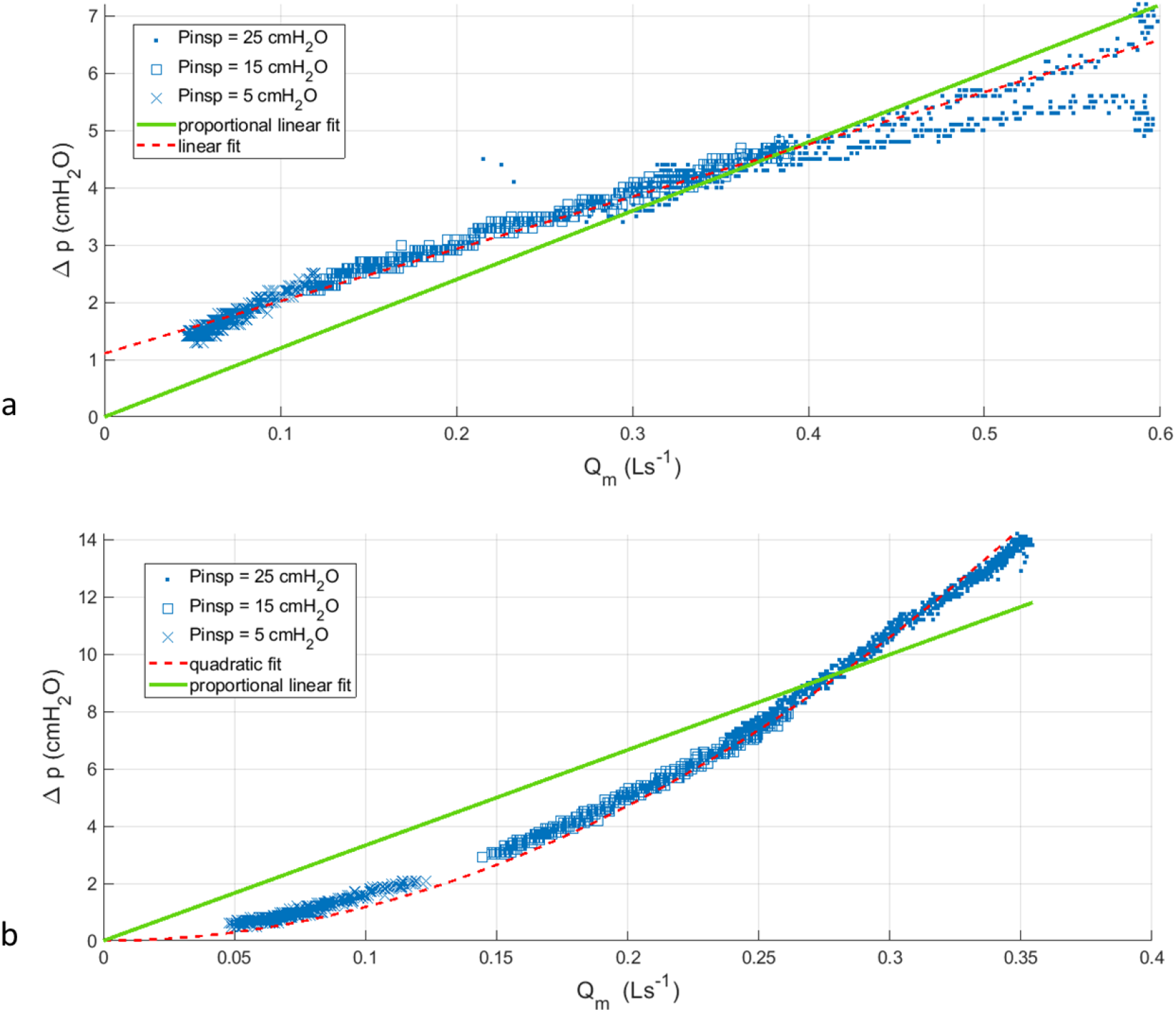
Characterisation plots from testing data using Circuit 1, a. is for the non-return valve, b is for the SOR device.

**Table 1.**
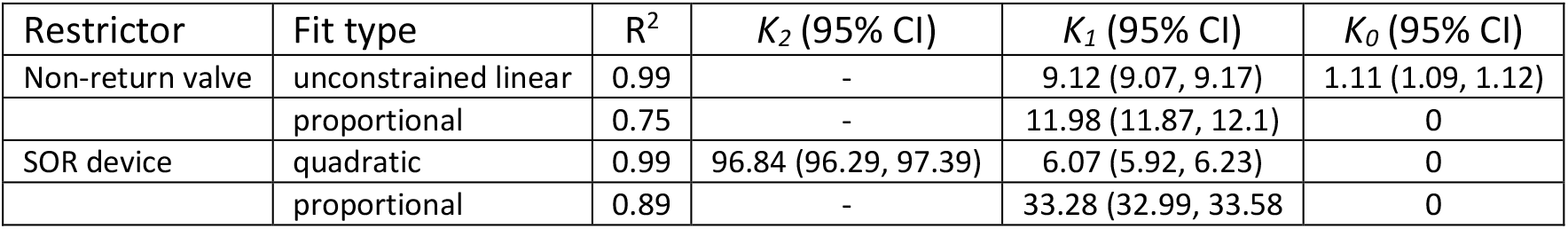
Fitting results for the two types of restrictor, for Pinsp values of 5, 15 and 25 cmH_2_O, with PEEP=5 cmH_2_O, RR=15 breaths/min, and I:E ratio is 1:2 for all tests

For parameter estimation (Circuit 2), *R* and *C* values for Lung 1 were 12 cmH_2_O/(L/s) and 0.040 L/cmH_2_O, respectively. For Lung 2, *R* and *C* values were 10 cmH_2_O/(L/s) and 0.030 L/cmH_2_O, respectively. The ventilator tubing resistance (*R_v_*) and compliance (*C_v_*) values were 22 cmH_2_O/(L/s) and 0.004 L/cmH_2_O respectively.

### Validation of BathRC Model

For validation test 1 (Figure 4 a, top row), without any additional resistance in the inspiratory limb for Lung 1, the flow was quite similar between the two test lungs. However, the higher compliance of Lung 1 was reflected in higher peak flow values. For both test lungs the BathRC model predictions for lung volume change were similar to the measured data (Figure 4 a, middle and bottom rows).

**Figure 4.**
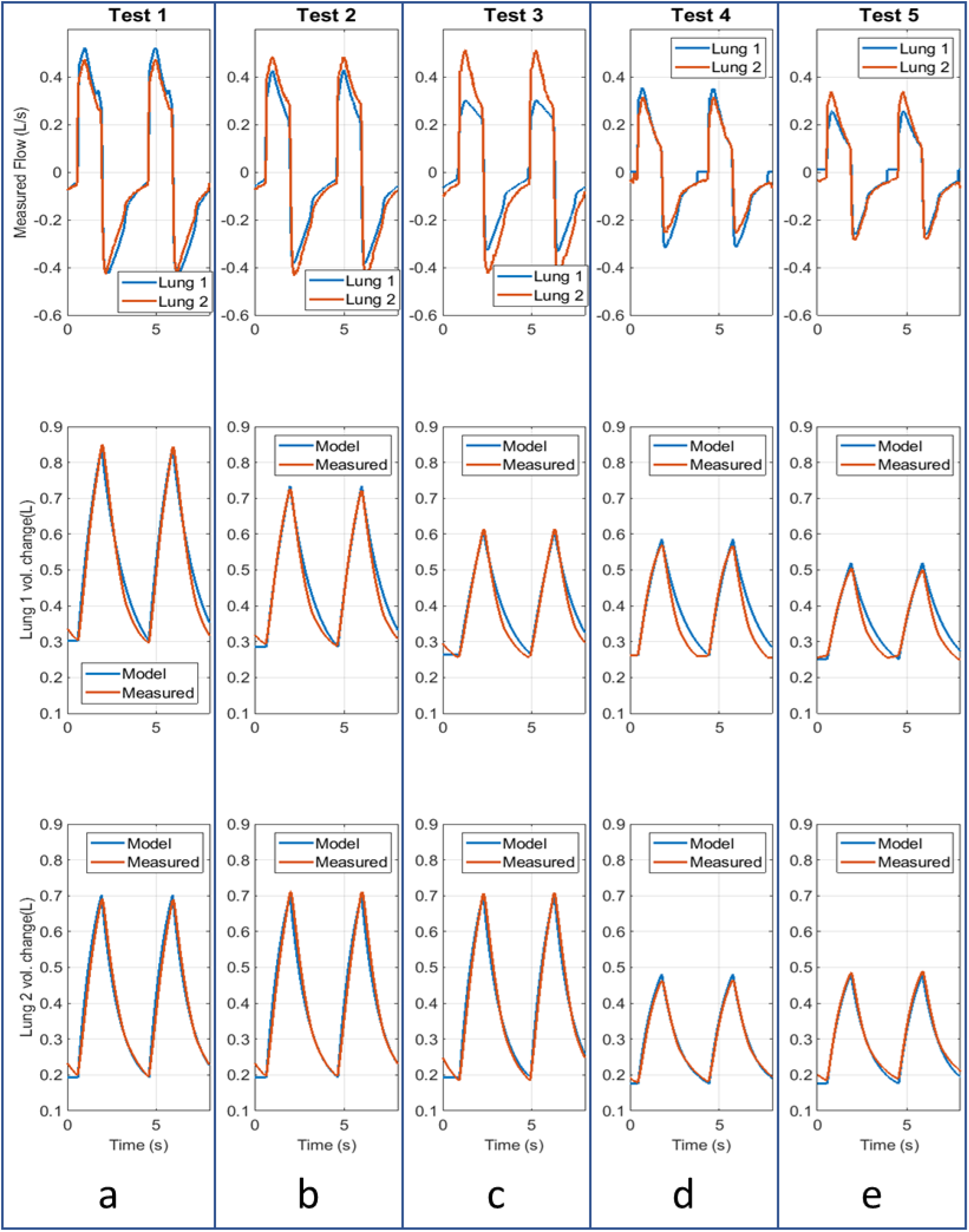
Plots of measured data, together with model prediction for the validation tests made using Circuit 3. a. Test 1, b. Test 2, c. Test 3, d. Test 4, e. Test 5.

For validation test 2, adding the non-return valve to the inspiratory limb for Lung 1 caused the peak flows for Lung 1 to be lower than those for Lung 2 (Figure 4 b, top row). Consequently, the tidal volume was reduced for Lung 1 (Figure 4 b, middle row), with model predictions again closely tracking the measured data (Figure 4 b, middle and bottom rows). The tidal volume for Lung 1 was reduced by 22% relative to its unrestricted state.

In validation test 3, restriction in the form of the SOR device further reduced the peak flow values for Lung 1 (Figure 4 c, top row). For Lung 1, the tidal volume reduction was 36% from its unrestricted state. For Tests 1 to 3, there was no change in the parameters for Lung 2, so the model predicted the same tidal volume in each case.

Similarly, when operating at 15 cmH_2_O Pinsp for validation tests 4 (Figure 4 d, top row) and 5 (Figure 4 e, top row), the effect of adding the non-return valve to provide restriction in test 5 is clearly seen. In both cases the model predictions matched the measured data well. Lung 1 tidal volume reduces by 20% as a result of the restriction, again associated with a small increase in Lung 2 tidal volume (6%).

The measured and predicted values for tidal volumes are given in Table 2 for the validation tests. The RC model predicts the tidal volumes reasonably well for this range of conditions. For Lung 1, in the five validation tests the measured tidal volume deviated from the predicted by 2.4%, −3.6%, 3.5%, −5.6% and−9.3%, respectively. For Lung 2, the measured tidal volume deviated from the predicted by −2.2%, 1.2%, 2.6%, −7.5% and −2.0% respectively. The largest absolute error was 25 mL.

**Table 2.** Test conditions, with predicted (from BathRC model) and measured tidal volumes, *V_T_*. PEEP=5 cmH_2_O, RR=15 breaths/min, and I:E ratio was 1:2 for all tests

| Test condition |  |  | Tidal volume (L) |  |  |  |
| --- | --- | --- | --- | --- | --- | --- |
| Validation test no. & related fig. | P<sub>insp</sub> (cmH<sub>2</sub>O) | Restrictor (Lung 1) | Lung 1 |  | Lung 2 |  |
|  |  |  | Predicted | Measured (stdev*) | Predicted | Measured (stdev*) |
| 1) Fig.4a | 25 | None | 0.541 | 0.554 (0.001) | 0.509 | 0.498 (0.005) |
| 2) Fig.4b | 25 | Non-return valve<sup>1</sup> | 0.449 | 0.433 (0.002) | 0.509 | 0.515 (0.004) |
| 3) Fig.4c | 25 | SOR<sup>2</sup> | 0.345 | 0.357 (0.004) | 0.509 | 0.522 (0.001) |
| 4) Fig.4d | 15 | None | 0.324 | 0.306 (0.007) | 0.305 | 0.282 (0.006) |
| 5) Fig.4e | 15 | Non-return valve<sup>1</sup> | 0.270 | 0.245 (0.005) | 0.305 | 0.299 (0.006) |

<sup>1</sup> Resistance 12 cmH<sub>2</sub>O/(L/s) <sup>2</sup> Resistance 33 cmH<sub>2</sub>O/(L/s)
\*stdev=standard deviation in measured tidal volume over 15 cycles

## DISCUSSION

To succeed with DPV, we believe it is essential to independently control the gas flow (tidal volume that each patient receives. A possible solution is to use a flow restrictor in the line of a patient who would otherwise receive too much pressure or flow, resulting in barotrauma or volutrauma. Such a restrictor should ideally be adjustable. We have presented experimental results using a pair of test lungs showing that restricting the flow in one inspiration line does indeed reduce the tidal volume in the corresponding lung. Moreover, using a linear resistance-compliance network model, we have shown that the change of tidal volume can be predicted. In the five tests presented, the largest prediction error was 25 mL of tidal volume. In the form used, the model just needs an airway resistance and lung compliance estimate for each patient, which is routinely available in a clinical setting, and a resistance and compliance value for the ventilator tubing system. Likewise, an added flow restrictor should be characterised by a linear resistance i.e. a pressure drop proportional to flowrate. The predictions of the model are good despite clear linearization errors for the two flow restrictors used in this study. All the parameter values used have been informed by individual component testing.

Some further observations on the results:

1. Non-return valves are used in the individual inspiratory and expiratory limbs of each circle system (four in total). The valves we used have a considerable resistance, 12 cmH_2_0/(L/s), so contributing over half of the total flow path resistance (either inspiration or expiration) estimated to be 22 cmH_2_0/(L/s). While this will mean the characteristics of the dual arrangement are markedly different from conventional single patient ventilation, the increased pressure loss within the flow path means that the airway flow and pressure will be less sensitive to changes in patient characteristics.
2. A result that was not predicted by the simple modelling was that as the flow reduced to one test lung by the introduction of a flow restrictor, there was a small increase in flow to the other lung. The most severe flow restrictor reduced tidal volume by 36% in the corresponding lung, but also increased the tidal volume by 5% in the unaltered loop for the other lung.

A key advantage of the BathRC model is that it is simple to implement and does not require iterative methods. As such, it can be straightforwardly implemented in a spreadsheet (supplementary information Appendix B), or as an online calculator. This allows clinicians to estimate the flow restriction needed to match patient requirements. The challenge remains, however, to source a flow restrictor which is clinically acceptable, and ideally adjustable. The 3D printed designs that are emerging need to be proven to be inert, sterilisable, and durable in the breathing system environment. The fixed restrictor used in this study – in fact a non-return valve – is clinically approved and provided around 10% differentiation between the two loops. Two or more could be used in series to provide a greater restriction, but an adjustable flow restrictor would be far easier to use and would limit the need to break the closed system to add additional resistance should it be required.

Some other issues which should be investigated are:

1. The effectiveness of the non-return valves in preventing retrograde flows between patients, and in ensuring unidirectional flow around each circuit, contributing to expired CO_2_ removal
2. The addition of sensors to give immediate feedback of the effect of flow restriction
3. The ability of a ventilator to maintain the specified pressure when the flow demands have doubled due to dual ventilation needs to be assured, as is the effectiveness of the CO_2_ absorber, although it is anticipated that high gas flows will be used

To reiterate, the APSF have recently recommended that ventilator sharing should not be undertaken^3^. Some of the objections raised are addressed in this work. We recognise that no-one would choose to share a ventilator between two patients, but there may be some situations when there will be no choice. We also recognise the additional challenge this set-up will present to those caring for patients in these circumstances. We believe that manageability and safety mandates limiting the sharing to two patients and not more. Dual patient ventilation is a method of last resort.

## Data Availability

BathRC model is available here for general usage.

https://github.com/arp23/BathRC

## ACKNOWLEDGEMENTS

We thank BMI Bath Clinic for providing test facilities. We also wish the acknowledge help and advice from the following University of Bath academics: Mauro Carnevale, David Cleaver, Andrew Cookson, Kate Fraser, Pejman Iravani, Evros Loukaides, Alexander Lunt, Anna Young and other members of the Bath Mechanical Ventilation Group.

## AUTHOR CONTRIBUTIONS

Andrew Plummer: concept, methodology, experimental planning, analysis, model creation, writing and reviewing.

Jonathan L. du Bois: concept, methodology, model creation, data analysis, writing, reviewing

Joseph Flynn: Design of SOM restrictor, CAD modelling, concept, writing, reviewing

Jens Roesner: Methodology, model creation, data analysis, writing, reviewing

Siu Man Lee: Planning, data collection, writing, reviewing

Patrick Magee: concept, planning, methodology, data collection, writing, reviewing

Malcolm Thornton: concept, planning, methodology, writing, reviewing

Andrew Padkin: planning, data collection, writing, reviewing

Harinderjit S. Gill: concept, planning, methodology, data collection, writing, reviewing

## DATA STATEMENT

The data are available from the University of Bath data repository.

## APPENDIX A

As shown in Figure 1, the model consists of the patient lung/airway resistance (*R*) and compliance (*C*), and a resistance (*R_v_*) and compliance (*C_v_*) representing the properties of the ventilator tubing system. The airway flow rate, q, is related to the rate of change in airway pressure *p_p_*, according to:

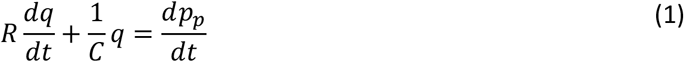

And flow from the ventilator, given by tubing pressure drop divided by resistance, is split between the patient and what is absorbed by tube compliance (including gas compressibility):

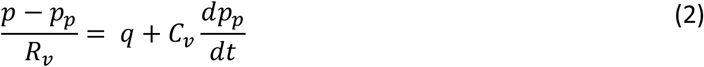

Combining equations (1) and (2), the gas volume reaching the patient’s lungs, v, can be related to the ventilator pressure, p, by:

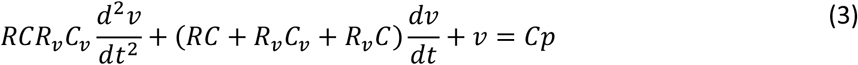

Physically realistic parameter values lead to a very small second derivative term, so:

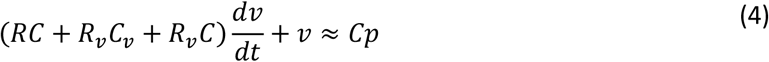

Due to ventilator or patient properties, the inspiration and expiration parameters may be different. Therefore, the first order response represented by (2) may have different inspiration and expiration time constants, given by:

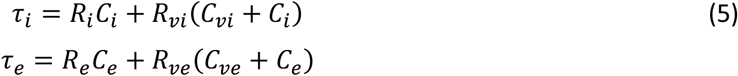

In pressure control, the ventilator pressure *p* approximates a series of square pulses. The lung volume during inspiration at time *t*, in addition to the Functional Residual Capacity (FRC) of the patient’s lungs, is given by:

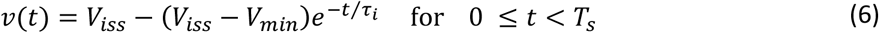

where *V_iss_* is the steady state asymptote of the inspiration phase, *V_min_* is the minimum volume, which occurs at the start of inspiration, and *T_s_* is the time at which the ventilator switches from high to low pressure (Figure A1). During expiration the volume is given by:

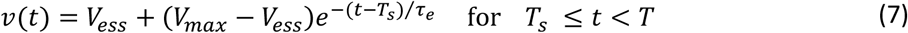

where *V_ess_* is the steady state asymptote of the expiration phase, *V_max_* is the maximum volume, which occurs at the start of expiration, and *T* is the period for the full breathing cycle. These functions are illustrated in Figure A1.

The steady state volumes are given by the product of compliance and pressure:

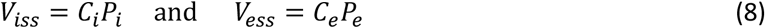

where *P_e_* is the low pressure setting of the ventilator (PEEP), and *P_i_*, is the high pressure used for inspiration.

Maximum volume is reach at the end of inspiration, i.e. at *t = T_s_*, and minimum volume is reached at the end of expiration, i.e. at *t = T*, so (6) and (7) can be written at these particular times as:

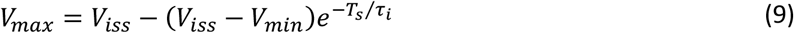

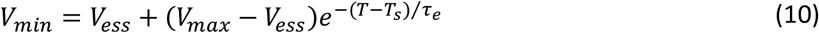

Define the following coefficients:

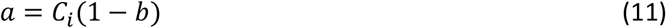

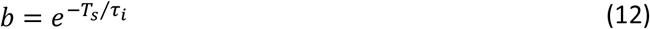

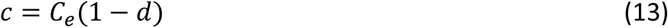

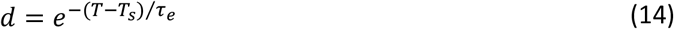

**Figure A1.**
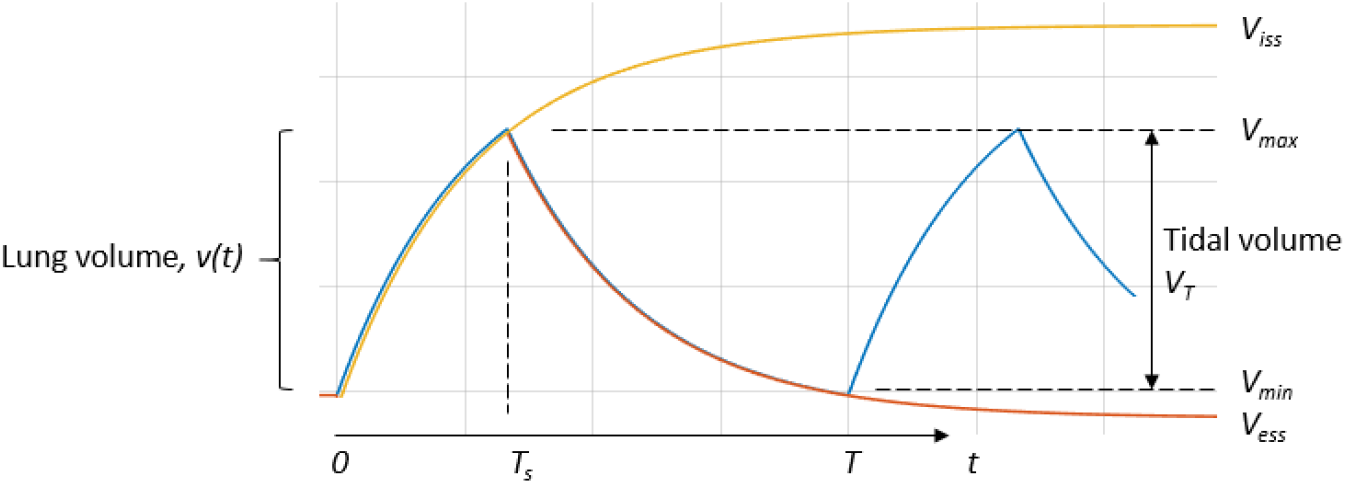
Lung volume waveform variable definitions

Thus the minimum and maximum volumes are related to the ventilator pressures by:

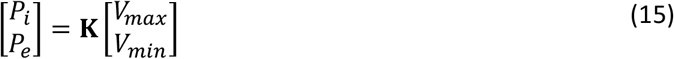

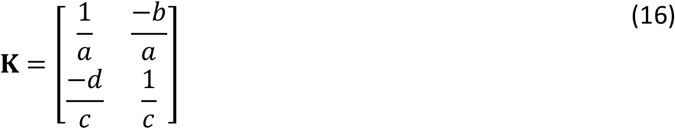

So the maximum and minimum volume and tidal volume (*V_T_*) can be calculated:

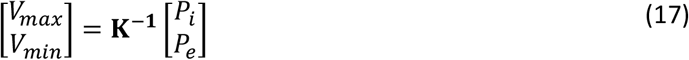

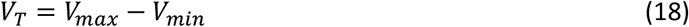

Equation (17) is equivalent to:

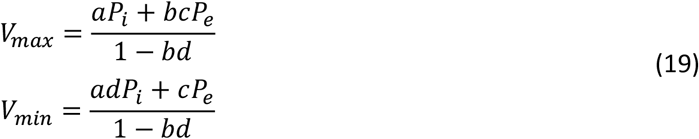

### Calculating inspiration restrictor resistance

The resistance required to achieve a specified tidal volume *Ṽ_T_* can be found. From (10), (14), and (18):

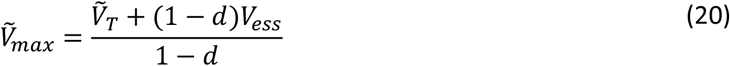

Hence *Ṽ_min_* can be found from (18), and the new inspiration time constant can be found from (9):

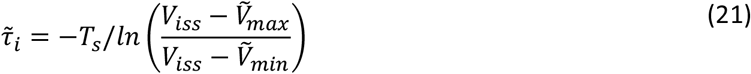

From (5), the additional resistance, *R_r_*, in the inspiration line can be found:

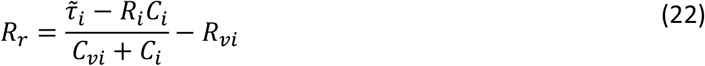

Alternatively, if the original tidal volume is required to be maintained but with an increased ventilator pressure *P_i_*, equation (21) can be used with a new value for the end-inspiratory steady state volume *V_iss_* given by (8), and (22) can be again used to find the restrictor resistance for the new inspiratory time constant.

As this model does not need a time-marching numerical solution, results can be determined by calculations in, for example, a spreadsheet. A view of such an implementation is given in Appendix B.

## APPENDIX B

A snapshot of a flow restrictor sizing spreadsheet

| Calculation of inspiration restrictor resistance to alter patient ventilation characteristics |  |  |  |  |
| --- | --- | --- | --- | --- |
| Of use for ventilator sharing between patients |  | White fields are for data entry |  |  |
| **Ventilator settings. Ventilator must in Pressure Control mode** |  |  | **Derived ventilator parameters** |  |
| Respiration rate (breaths / min) | 15.0 |  | Respiration period (s) | 4 |
| Inspiration to expiration time ratio (I/E) | 0.5 |  | I to E switching time within period (s) | 1.33 |
| Pinsp (inspiration pressure above PEEP, cmH2O) | 25.0 |  | Max ventilator pressure (cmH2O) | 30.00 |
| PEEP (cmH2O) | 5.0 |  |  |  |
| Tubing system resistance (cmH2O/(L/s) ) | 20.0 |  |  |  |
| Tubing system compliance (L/cmH2O) | 0.0060 |  |  |  |
| **Patient characteristics** | A | B | C | D |
| Inspiration resistance, Ri (cmH2O/(L/s) ) | 13.0 | 13.0 | 13.0 | 13.0 |
| Expiration resistance, Re (cmH2O/(L/s) ) | 13.0 | 13.0 | 13.0 | 13.0 |
| Inspiration Compliance, Ci (L/cmH2O) | 0.054 | 0.044 | 0.034 | 0.024 |
| Expiration Compliance, Ce (L/cmH2O) | 0.054 | 0.044 | 0.034 | 0.024 |
| **Derived patient parameters** |  |  |  |  |
| Patient inspiration time constant, RiCi (s) | 0.70 | 0.57 | 0.44 | 0.31 |
| Patient expiration time constant, ReCe (s) | 0.70 | 0.57 | 0.44 | 0.31 |
| **Derived system parameters** |  |  |  |  |
| Ventilation inspiration time constant (s) | 1.90 | 1.57 | 1.24 | 0.91 |
| Ventilation expiration time constant (s) | 1.90 | 1.57 | 1.24 | 0.91 |
| Steady end-inspiration lung volume (L) | 1.62 | 1.32 | 1.02 | 0.72 |
| Steady end-expiration lung volume (L) | 0.27 | 0.22 | 0.17 | 0.12 |
| Max lung volume, in addition to FRC (L) | 1.045 | 0.903 | 0.753 | 0.587 |
| Min lung volume, in addition to FRC (L) | 0.461 | 0.345 | 0.238 | 0.145 |
| **Tidal volume (L)** | **0.584** | **0.557** | **0.515** | **0.442** |
| **Inspiration restriction to achieve a specified tidal volume** |  |  |  |  |
| Required tidal volume (L). Must be <= value above | 0.442 | 0.442 | 0.442 | 0.442 |
| New max lung volume, in addition to FRC (L) | 0.856 | 0.761 | 0.670 | 0.587 |
| New min lung volume, in addition to FRC (L) | 0.414 | 0.319 | 0.228 | 0.145 |
| New ventilation inspiration time constant (s) | 2.925 | 2.292 | 1.634 | 0.913 |
| **Inspiration restrictor needed (cmH2O/(L/s))** | **17.04** | **14.39** | **9.81** | **0.03** |
| OR inspiration added compliance needed (L/cmH2O) | 0.051 | 0.036 | 0.020 | 0.000 |
| **Inspiration restriction to achieve the same tidal volume (row 32) with a higher Pinsp pressure** |  |  |  |  |
| Higher Pinsp (insp. pressure above PEEP, cmH2O) | 30.00 | 30.00 | 30.00 | 30.00 |
| New steady end-inspiration lung volume (L) | 1.89 | 1.54 | 1.19 | 0.84 |
| New ventilation inspiration time constant (s) | 2.537 | 2.122 | 1.714 | 1.321 |
| **Inspiration restrictor needed (cmH2O/(L/s))** | **10.59** | **11.00** | **11.80** | **13.63** |

Note: 13 is median for normal lung, Arnal 2018
Note: same as inspiration often used
Note: 0.054 is median for normal lung, Arnal 2018
Note: same as inspiration often used
One breathing cycle, no restrictor
Note: 13 is median for normal lung, Arnal 2018 Note: same as inspiration often used Note: 0.054 is median for normal lung, Arnal 2018 Note: same as inspiration often used

